# Modeling the USA Winter 2021 Update and Spring 2022 CoVID-19 Resurgence

**DOI:** 10.1101/2022.06.24.22276889

**Authors:** Genghmun Eng

## Abstract

Every USA wave, including this latest Spring 2022 Resurgence has been successfully modeled using:

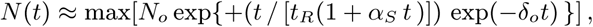

with *N* (*t*) being the total number of new CoVID-19 cases above a prior baseline, and *t*_*R*_ setting the pandemic wave doubling time *t*_*dbl*_ = *t*_*R*_ (In 2). The parameters {*α*_*S*_ ; *δ*_*o*_} measure mitigation efforts among the *uninfected population*. Here, {*α*_*S*_ *>* 0} is associated with *Social Distancing* and *vaccinations* ; while {*δ*_*o*_ *>* 0} is associated with *mask-wearing*, which results in faster 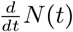 post-peak *drop-offs*. The predicted pandemic wave end is when the calculated *N* (*t*) stops increasing.

The USA Winter 2021 Resurgence resulted in fewer *Omicron* CoVID-19 cases than calculated in our prior *medrxiv*.*org* preprint^*^, due to an increased *δ*_*o*_ component, which gives ∼3*/*11*/*22 as the predicted wave end. A relatively quiet CoVID-19 period ensued until ∼4*/*16*/*22, when a new *Omicron* variant caused the present Spring 2022 CoVID-19 resurgence.

The recent CoVID-19 waves have decreasing *t*_*R*_ values, with the Spring 2022 *t*_*R*_ *≈* 3.55 *days* (*t*_*dbl*_ ≈ 2.46 *days*) value being the shortest since the initial 2020 pandemic, indicating increasingly infectious variants. The Winter 2021 and the present Spring 2022 CoVID-19 resurgences have identical *α*_*S*_ ≈ 0.043 */ day* values, but the *δ*_*o*_ *>* 0 *mask-wearing* parameter decreased from *δ*_*o*_ ≈ 3. 14 × 10^−3^*/ day* to *δ*_*o*_ ≈ 1. 145 × 10^−3^*/ day*, giving the Spring 2022 wave a longer tail, and an expected end date of 8/25/22, with these wave totals:

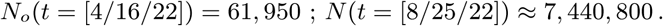

When all the USA CoVID-19 waves are combined, it gives:

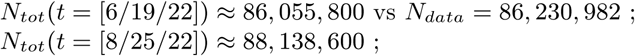

assuming no future CoVID-19 Resurgences (*with 5 Figures*).

*\*(10*.*1101_2022*.*02*.*04*.*22270491)*

## 1 Introduction

Each USA CoVID-19 wave^**1–8**^, from the pandemic start (3/21/20) to the present day (6/19/22), has been successfully modeled using the following basic *N* (*t*) function for the total number of new CoVID-19 cases above a prior baseline:

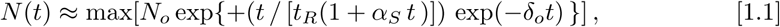

with *t*_*R*_ setting the pandemic doubling time *t*_*dbl*_ = *t*_*R*_ (ln 2), as in standard **SEIR** (**S**usceptible, **E**xposed, **I**nfected, **R**ecovered or **R**emoved) epidemiology models. Mitigation efforts among the *uninfected population* in Eq. [1.1] has *α*_*S*_ *>* 0 associated with *Social Distancing* and *vaccinations*; and *δ*_*o*_ *>* 0 associated with *mask-wearing*, which results in a faster 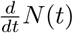 post-peak *drop-off*. The predicted pandemic wave end is when the calculated *N* (*t*) stops increasing.

Given a total population of *N*_*ALL*_, the *uninfected population U* (*t*) is:

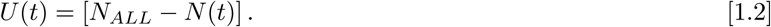

Using Eq. [1.1] assumes *N* (*t*) *<< N*_*ALL*_, so that pandemic saturation effects can be ignored. Also, **SEIR** models do not generally include what the *U* (*t*) *uninfected population* is doing in response to the pandemic. In contrast, Eq. [1.1] was developed as a *non-local* extension of **SEIR** models, to account for how the *uninfected population*, as a whole, is mitigating the pandemic spread.

Each new USA CoVID-19 wave starts with a sharp rise in the total number *N* (*t*) of new cases, with the *t* = 0 point for each wave being determined by when the resurgence is first easily identified, where *N* (*t* = 0) = *N*_*o*_ is the number of cases above baseline at that time. Although Eq. [1.1] does not predict when each new CoVID-19 wave starts, or what conditions are causing the new wave, once the CoVID-19 wave becomes established, Eq. [1.1] appears to successfully predict its time evolution.

The fact that the same few parameters in Eq. [1.1] have successfully modeled the time evolution of each USA CoVID-19 wave^**1–8**^, shows that the **response** of the *U* (*t*) *uninfected population* has been similar for each wave, even though different factors may have been driving each CoVID-19 resurgence. All CoVID-19 data used here came from the open-source *bing*.*com* CoVID-Tracker^**9**^ database.

## 2 Winter 2021 / Spring 2022 CoVID-19 Update

Our prior *medrxiv*.*org* preprint^**8**^ for the Winter 2021 resurgence showed an *initial* stage (11/15/21-12/25/21) which had practically no *mask-wearing* effects [*δ*_*o*_ *<* 0 001 × 10^−3^*/ day*], likely due to the *Omicron* variant infecting vaccinated people who thought they were protected. It was followed by a *latter* post-Christmas stage (12/25/21-1/31/22), having these parameter values:

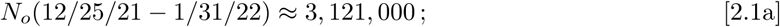

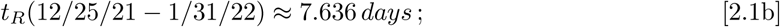

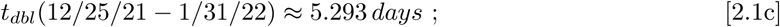

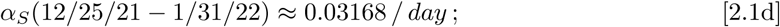

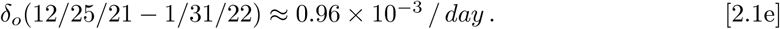

The *N*_*o*_(12/25/21) = 3, 121, 000 starting point for a behavior change in *N* (*t*) was similar to the *latter* stage of the Summer 2021 Resurgence^**6**^, which also showed a behavior change in *N* (*t*) starting at *N*_*o*_(8/13/21) = 3, 200,000. This commonality shows that the *uninfected population* altered its behavior at similar points, which is likely when people realized that some hospitals would be overwhelmed.

A further update for the US Winter 2021 CoVID-19 Resurgence is presented here in *Figs. 1-2*, which shows that a significant increase in *mask-wearing* in early 2022 likely helped to curtail this CoVID-19 resurgence, making it less pernicious than Eqs. [2.1a]-[2.1e] initially indicated. The final fit for this *latter* part of the Winter 2021 resurgence has these updated values:

**Fig. 1:**
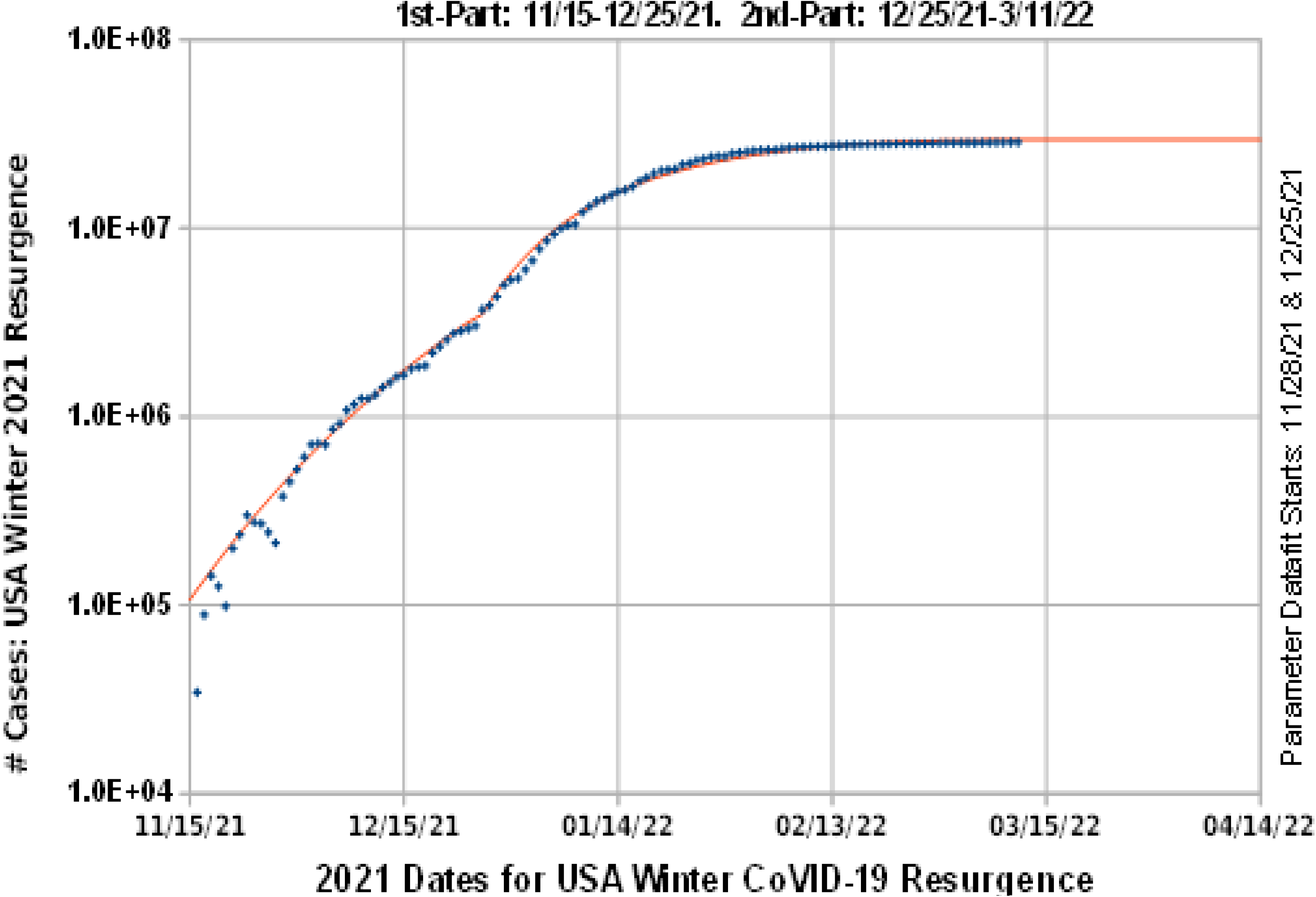
Final Fit, USA Winter 2021 Omicron CoVID-19 Wave by Itself

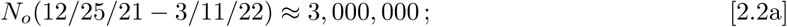

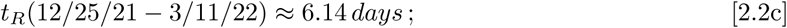

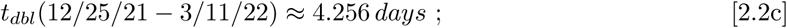

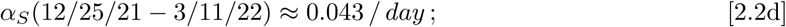

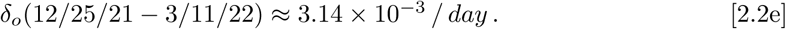

The major change was a more than three-fold increase in the critical *mask-wearing 8*_*o*_-parameter. This updated data and model for the US Winter 2021 CoVID-19 Resurgence also provides a better baseline for follow-on resurgences. While *Fig. 2* only presents the modeling results of Eq. [2.2a]-[2.2e] through 3/11/2022, it also includes data up through 6/19/2022. Those data show that the US is now in the midst of a US Spring 2022 CoVID-19 Resurgence, as new CoVID-19 variants continue to infect both vaccinated and unvaccinated people.

**Fig. 2:**
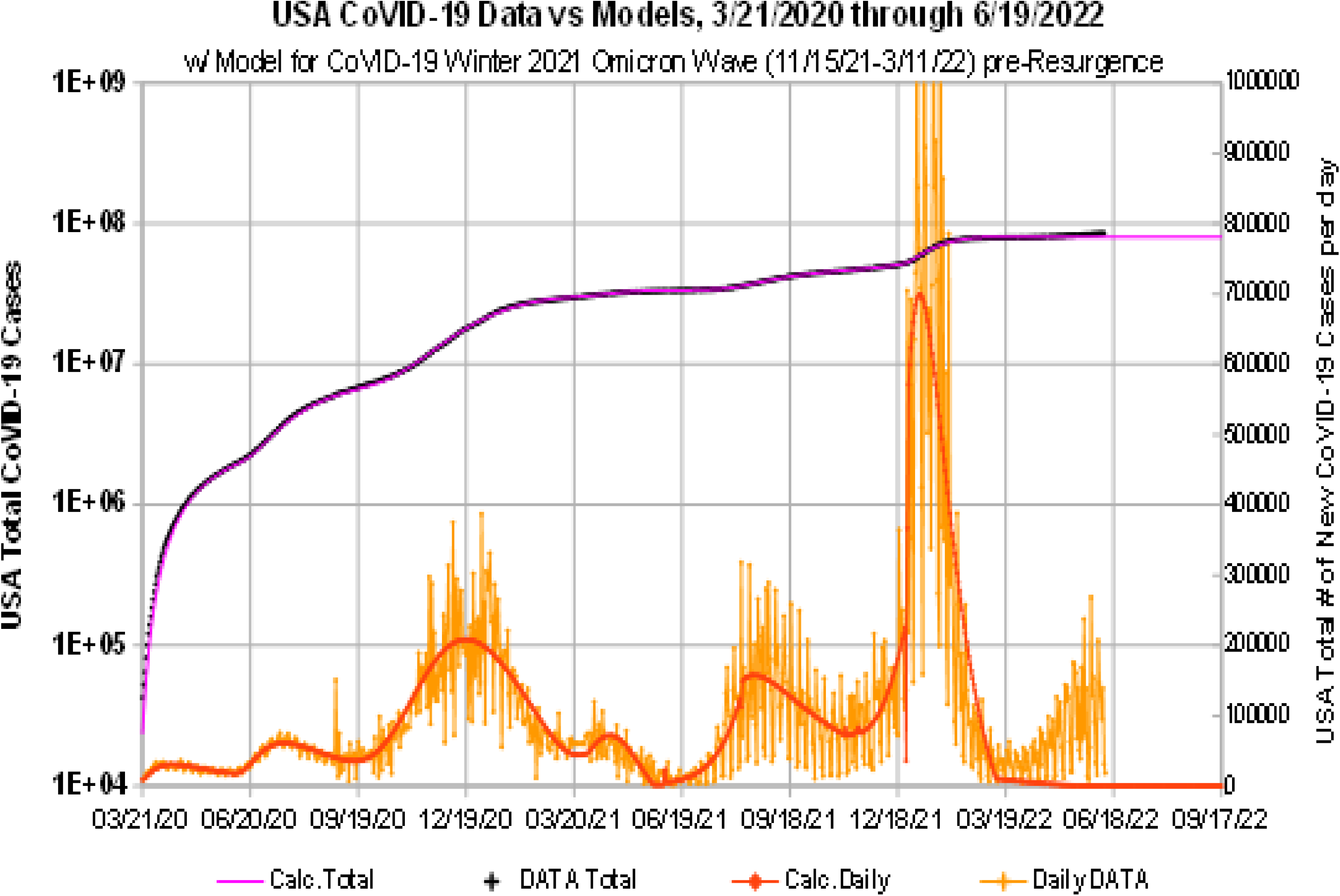
USA CoVID-19 Totals w/ Data Fitting 3/21/2020 - 3/11/2022

Using *Fig. 2* and Eqs. [2.2a]-[2.2e] as a new baseline then determines these parameters for the US Spring 2022 CoVID-19 Resurgence:

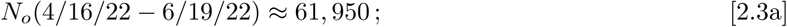

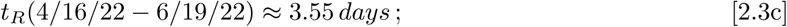

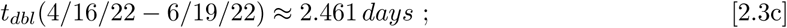

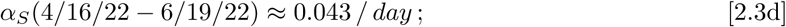

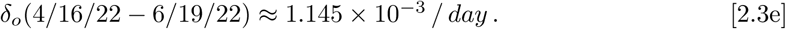

*Figure 3* compares the Eq. [2.3a]-[2.3e] predictions for the Spring 2022 resurgence to the data above baseline. The combined model for all the US CoVID-19 waves is shown in *Fig. 4*, going back to March 2020.

**Fig. 3:**
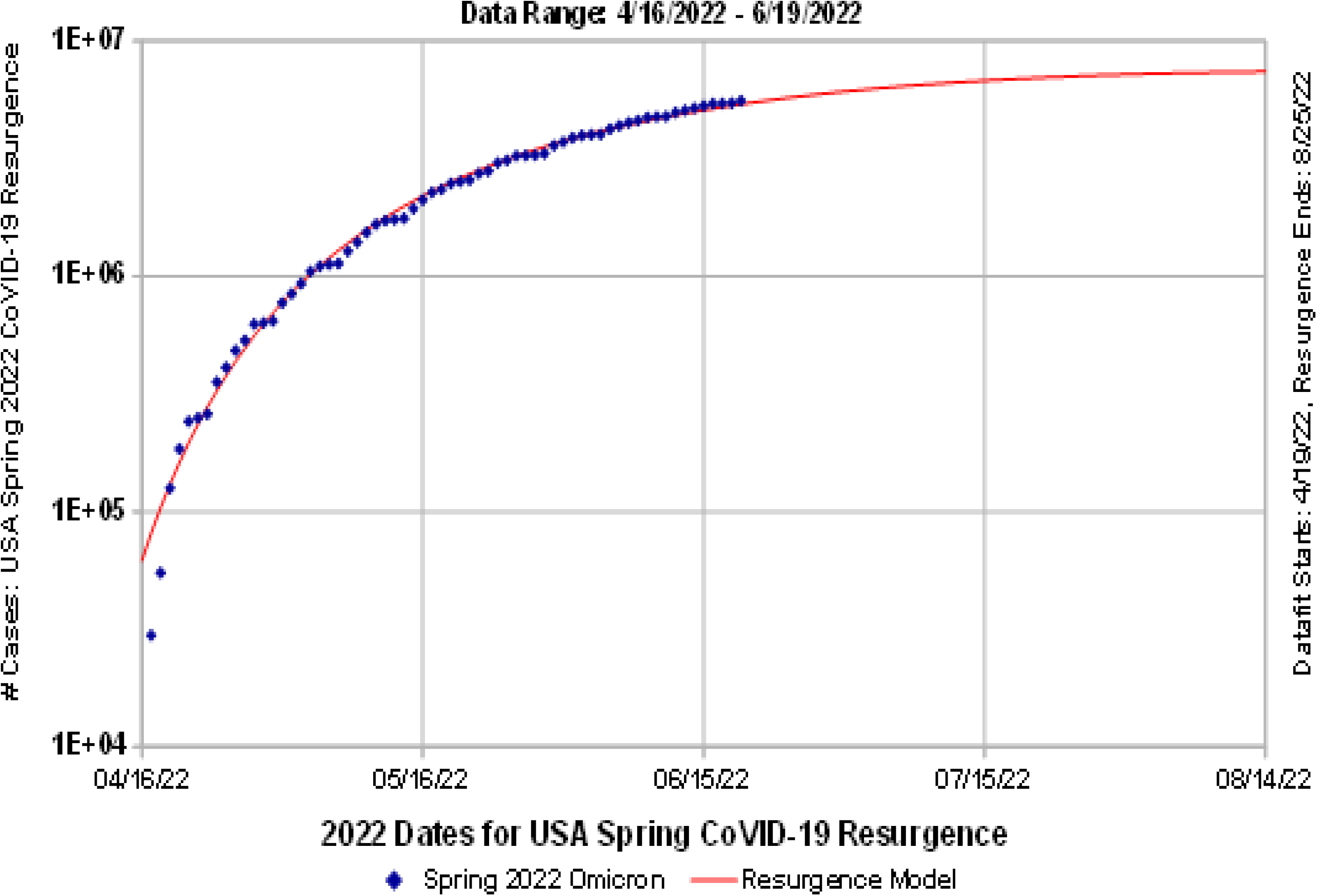
USA CoVID-19 Spring 2022 Resurgence By Itself

**Fig. 4:**
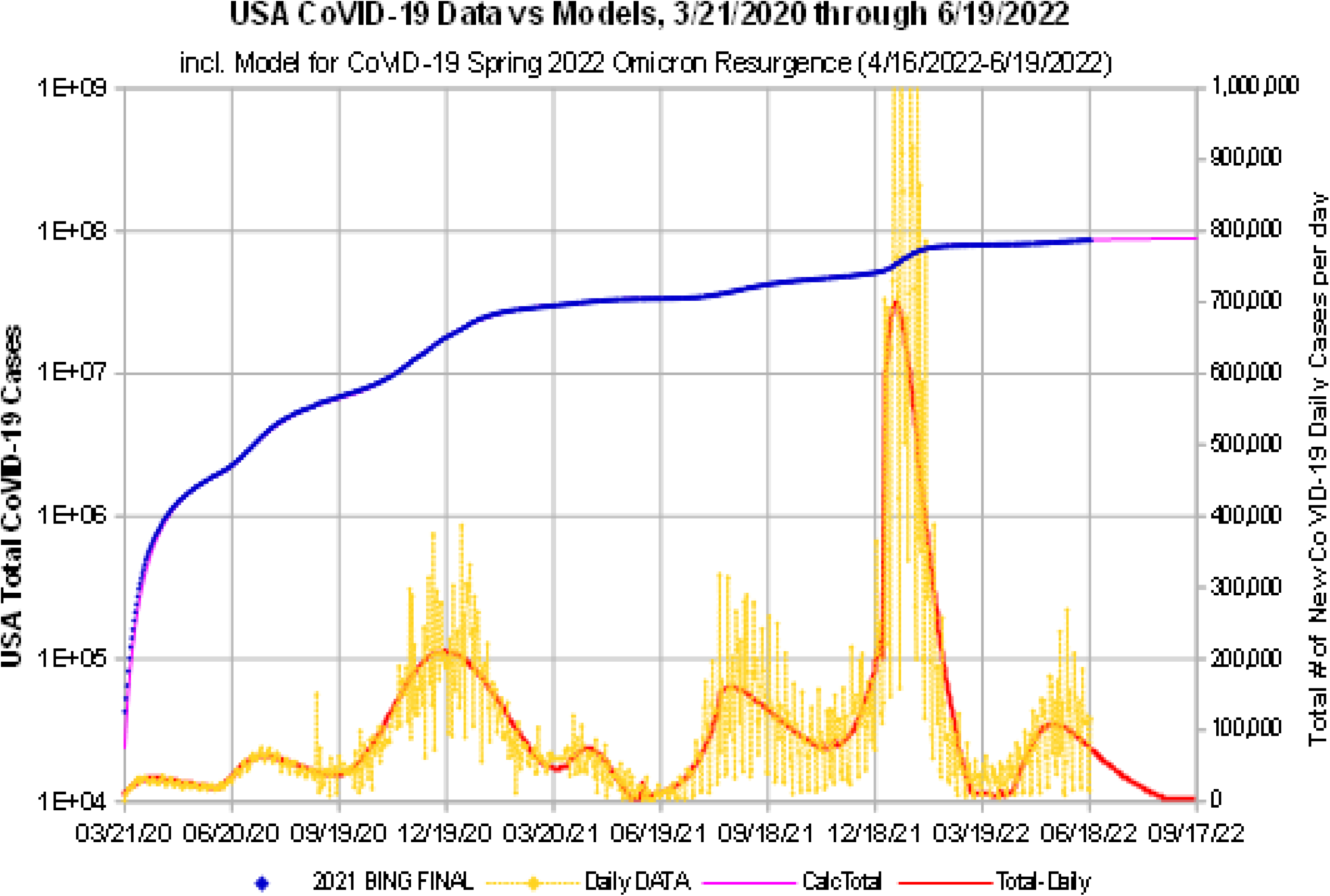
USA CoVID-19 Daily and Total, w/ Spring 2022 Resurgence

While Eq. [1.1] captures the overall structure of each new CoVID-19 wave, it does not successfully model the lowest values for the long-term tail of the daily CoVID-19 infections. It also does not predict when the next resurgence begins, since that *t* = 0 point is a model-adjusted input parameter determined by when the new resurgence is first easily identified. Both these aspects are apparent in the *Fig. 4* modeling gap between the 3/11/22 predicted end of the Winter 2021 Resurgence, and 4/16/22, when the Spring 2022 Resurgence reached its *N*_*o*_(4/16/22) ≈ 61, 950 initial modeling threshold. These results also mean that the US was nearly free of CoVID-19 between these two pandemic waves.:

## 3 Summary

Our empirical modeling of the various USA CoVID-19 pandemic waves shows that there is a fairly consistent range of responses by the *uninfected population* to the various CoVID-19 waves, even though each wave may have had a different dominant root-cause. *Figure 5* tabulates all the model parameters that were derived for each CoVID-19 wave.

**Fig. 5:**
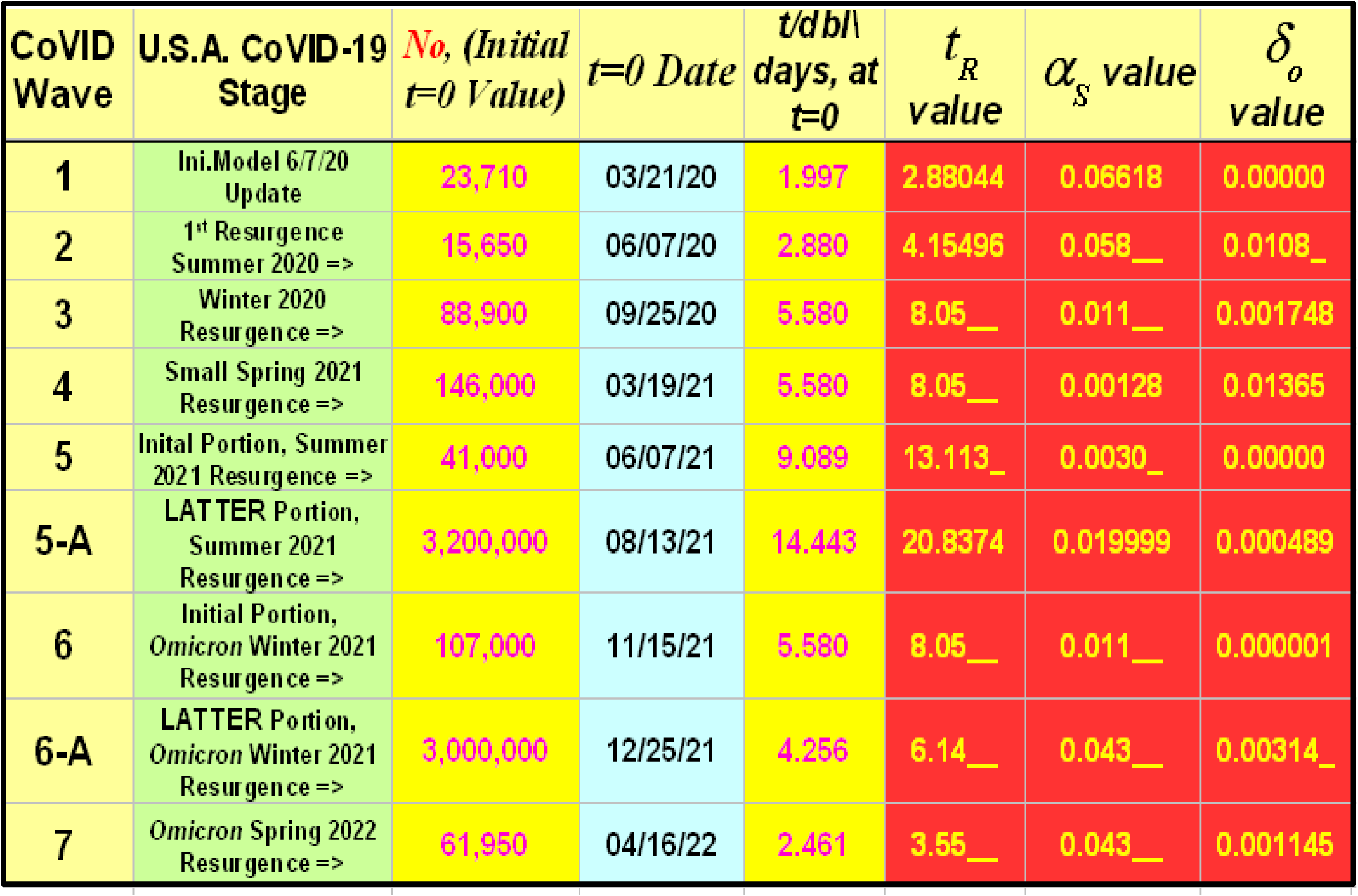
Summary of CoVID-19 Model and Parameter Values

It is now generally accepted that *Omicron* evades the CoVID-19 vaccines, but that vaccination still offers protection against hospitalization. Since this variant was detected in Nov. 2021, our modeling since then (*Fig. 5: CoVID Waves 6, 6-A*, and *7*) shows that each *Omicron* wave was intrinsically more infectious, with *t*_*dbl*_ values decreasing from ∼5 58 *days* down to its most recent value of ∼2 46 *days*, which is shorter than every wave, except for the initial March 2020 pandemic value of ∼2 00 *days*.

Because the initial *Omicron CoVID-19* wave caught the vaccinated population by surprise, it had a low *α* _*S*_ ≈ 0.011 */ day* value, with virtually no *mask-wearing* effects (*δ* _*o*_ ≲ 0.001 × 10^−3^ */ day*). However, the post-Christmas data showed a significant increase in *Social Distancing*, which gave *α* _*S*_ ≈ 0 043 */ day* for both the *latter* part of the Winter 2021 Resurgence, and for the present follow-on Spring 2022 Resurgence. The *δ*_*o*_ change from a post-Christmas value of *δ*_*o*_ ≈ 3.14 × 10^−3^ */ day*, to the present Spring 2022 lower value of *δ*_*o*_ ≈ 1.145 × 10^−3^ */ day*, means that this latest wave has a relatively long-tail for the CoVID-19 daily cases, which is also apparent in *Fig. 4*.

These analyses give the following updated USA CoVID-19 projections for the total number of USA CoVID-19 cases:

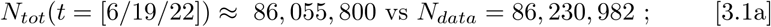

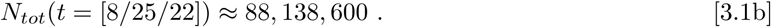

Unfortunately, the latest international CoVID-19 pandemic data indicates that newer vaccine-evading *Omicron* variants, which are not included in this model, are coming to the USA. Vigilance, *Social Distancing*, and *mask-wearing* will continue to be needed.

## Data Availability

All data produced in the present work are contained in the manuscript.

## Notes

### Competing Interest Statement

The authors have declared no competing interest.

### Funding Statement

This study did not receive any funding.

### Author Declarations

All CoVID-19 data used here came from the open-source bing.com CoVID-Tracker⁹ database: www.bing.com/covid/local/unitedstates?form=COVD07

